# LOCKDOWN AS A PANDEMIC MITIGATING POLICY INTERVENTION IN INDIA

**DOI:** 10.1101/2020.06.19.20134437

**Authors:** Subhayan Mandal, Manoj Kumar, Debasish Sarkar

**Affiliations:** Malaviya National Institute Of Technology, Jaipur, Rajasthan-302017, India

**Keywords:** Epidemiology, Covid-19, Variable Reproduction Number, Outbreak Policy Intervention

## Abstract

We use publicly available timeline data on the Covid-19 outbreak for nine indian states to calculate the important quantifier of the outbreak, the sought after *R*_*t*_ or the time varying reproduction number of the outbreak. This quantity can be measured in in several ways, e.g. by application of Stochastic compartmentalised SIR (DCM) model, Poissonian likelihood based (ML) model & the exponential growth rate (EGR) model. The third one is known as the effective reproduction number of an outbreak. Here we use, mostly, the second one. It is known as the instantaneous reproduction number for an outbreak. This number can faithfully tell us the success of lockdown measures inside indian states, as containment policy for the spread of Covid-19 viral disease. This can also, indirectly yield notional value of the generation time inteval in different states. In doing this work we employ, pan India serial interval of the outbreak estimated directly from data from January 30^th^ to April 19^th^, 2020. Simultaneously, in conjunction with the serial interval data, our result is derived from incidences data between March 14^th^, 2020 to June 1^st^, 2020, for the said states. We find the lockdown had marked positive effect on the nature of time dependent reproduction number in most of the Indian states, barring a couple. The possible reason for such failures have been investigated.

## 1. Introduction

Global pandemic outbreaks are very common nowadays. India is no exception. The Severe Acute Respiratory Syndrome (2003), [1] Avian Influnza (2006), [2] Swine Flu (2015) [3] are to name a few. There were others that did not touch upon India but were recent events, such as MERS (2012) [4] & EVD (Ebola virus disease) (2000, 2003, 2004 & most recently in 2012) [5]. None of the above, however, touched the global pandemic scale, of what has been attained by Novel Coronavirus, aka Covid-19, in a short span of time, starting at the end of past year [6, 7, 8]. The global community responded to this unprecedented situation by various policy interventions. Wearing masks & face shields [9, 10] in public, social distancing norms [11] were amongst them. More drastic & perhaps draconian step of lockdown [12] was taken by governments across the world, as a containment policy measure [13]. We analyse the effect of lockdown on the propagation of Covid-19 viral disease. The instantaneous version of basic reproduction number [14] of the infection is plotted against time to gauge the success [15] (or lack thereof) [16] of this policy intervention in nine different states of India. In the following, it is shown that this pervasive containment policy has borne fruit in most of the considered provinces.

## 2. Instantaneous Reproduction number

Time dependent or instantaneous reproduction number [17] is an accurate projection is an in-situ description of virility or virulence of epidemic diseases. The basic reproduction number [14] gives us the average number of infectee cases per infector from the previous generation, over a given period of time, in a fully susceptible population. Various policy implimentation and containment measures appreciably reduce the number of contacts, in turn reducing, the effective number [18] of susceptible contacts per potential infector. Epidemiologist have devised a time dependent parameter, effective reproduction number to assimilate the effect of policy intervention into the basic reproduction number during an ongoing epidemic. This quantity is defined as follows: Consider an individual, who turns infectious on day *t*. We denote by R_e_(t) the expected number of secondary cases this infectious individual causes, in future [19]. The instantaneous reproduction number, on the other hand, compares the number of new infections on day *t* with the infection pressure (force of infection) [20] from the days prior to *t*. It can be interpreted as the average number of secondary cases that each symptomatic individual at time *t* would infect, if the conditions remained as they were at time *t*. Hence, the stepwise, undulations, crests, troughs & spikes of this estimate is termed as instantaneous or real time measures.

There are various ways to calculate this effective instantanous & other time varying reproduction number. They are such to be:

### 2.1. Stochastic dynamic contact model-based method [21]

A stochastic Susceptible-Infected-Removed (SIR) model is considered in this case, in place of a deterministic one. Stochastic dynamic model has advantages over the standard deterministic one, in that, it accomodates improved variabilities and allows for better quantification of uncertainies of that number as compared to the standard deterministic model. Here, *S*(*t*), *I*(*t*) & *R*(*t*) denote the number of susceptible, infectious and recovered population at time respectively, and that *N* = *S*(*t*) + *I*(*t*) + *R*(*t*) is the total population. The infectious period of an infected individual is a random variable *T* ∼ exp (*γ*) & the reproduction rate is 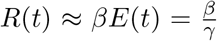, where *β* & *γ* are the transmission rate and recovery rate. The mathematical essence of the model can captured by a set of four coupled first order linear homogenous differential equations given such to be

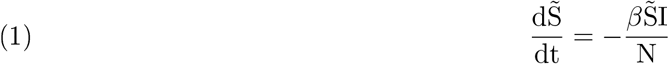

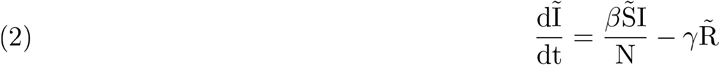

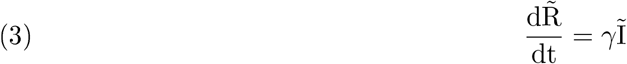

Here, ∼ signifies deterministic (average) counterparts. We set S (0) equals the population of the region, R(0) = 0, I (0) is 10 to 14 times the average number of confirmed cases from Day 0 to Day 7, and *γ* the inverse of mean infectious period, obtained from the parametrization of serial interval distribution collected directly from data described in section (3). The main difficulty with this time varying reproduction number is that it assumes a constant transmissibility, where it may vary & often peak, during the generation time interval and just before the onset of symptoms [20]. This model also can not accomodate various disease traits like asymptomaticity (non-detection), or human interferences like isolation measures or migration etc. Hence we do not look at this method any further here.

### 2.2. Poissonian likelihood based (ML) model [21]

Here it is assumed that the total number of secondary infectees that were infected by a single praimary infector follows a Poisson distribution. The number of individuals infected on (discrete) Date t is usually replaced by the number of daily incidences reported, on the same Date t. Also, the generation time interval is suitably replaced by the corresponding serial interval interval for all practical purposes. Let N_t_ be the number of reported incidences on Day t. Assuming that the serial interval has a maximum of k days and the number of new cases generated by an infected individual is assumed to follow a Poisson distribution with parameter [22] R. The probability that the serial interval of an individual lies in j days is w_j_, which can be estimated from the empirical distribution of serial interval or by setting up a discretized Gamma prior on it. Note only the nonnegative values of serial interval are used here. Thus, the likelihood function can be reduced into a thinned Poisson distribution as such

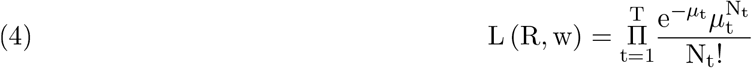

where,

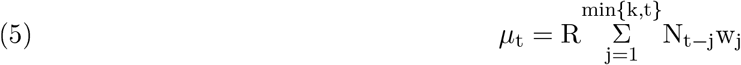

The instantaneous reproduction number can then be estimated by maximising the likelihood function as follows

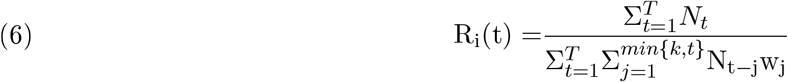

### 2.3. Exponential growth rate-based (EGR) method [21]

In the early days of the epidemic the number of infected cases rise exponentially. The growth rate (Malthusian coefficient) *r* can be estimated by fitting a non linear least square fitting into the daily incidence curve. The probability density function of serial interval of the outbreak is denoted by *f*_*λ*_(*t*), then the effective reproduction number is given by the Euler Lotka (type) Equation

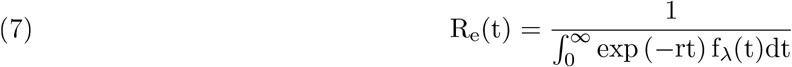

in case we have a non parametric serial interval distribution then we can define our effective reproduction number as

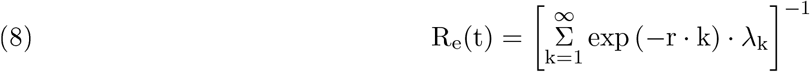

where *λ*_*i*_ are the observed serial intervals.

## 3. Serial Interval Distribution & Parametric Fit

Using the publically available data on github [23], to create a contact list between infector infectee pairs in the pan Indian context, between 30^th^ January-2020 to 19^th^April-2020. The data is then fitted with a log normal / gamma distribution to parametrize the values of mean and standard deviation.

## 4. Results

From the available data on github [23], the daily confirmed case incidences were collected for nine states, for the duration of 14^th^ March-2020 to 1^st^ June-2020. Applying the Poissonian ML method, the instantaneous R_i_(t) was plotted for each one of them, as given in figure number two to ten.

## 5. Discussion

It has been seen from above that, in all the state except Gujrat & Karnataka, the lockdown as a containment measure has been quite successful. In India the lockdown started from from 25^th^March-2020. After 11 to 13 days of the commencement of the same, seven provinces of India has shown us a significant downtrend of instantaneous reproduction number. This lag between cause & its effect corresponds to serial time or the generation time interval, that varies from state to state. However, these two states show a opposite trend. After the passage of about one generational time interval, the instantaneous reproduction number peaks sharply. This might correspond to migration [24] at the beginning of the lockdown. In Karnataka, however the value of instantaneous reproduction number fluctuates moderately. This is perhaps due to clustering [25] or inadequate testing policies [26], which may be true for both of the states. In what follows next are two province specific case studies.

## 6. Case Study

To be doubly sure, that our time dependent reproduction numbers [27] are calculated correctly over time, we shall fit the daily incidence graph of the two provinces which showed contrarian nature in reproduction numbers, during the initiation phase of lockdown. The time Dependent reproduction numbers shall be used to fit the daily incidences. The result is given in fig.(11).

**Figure 1.**
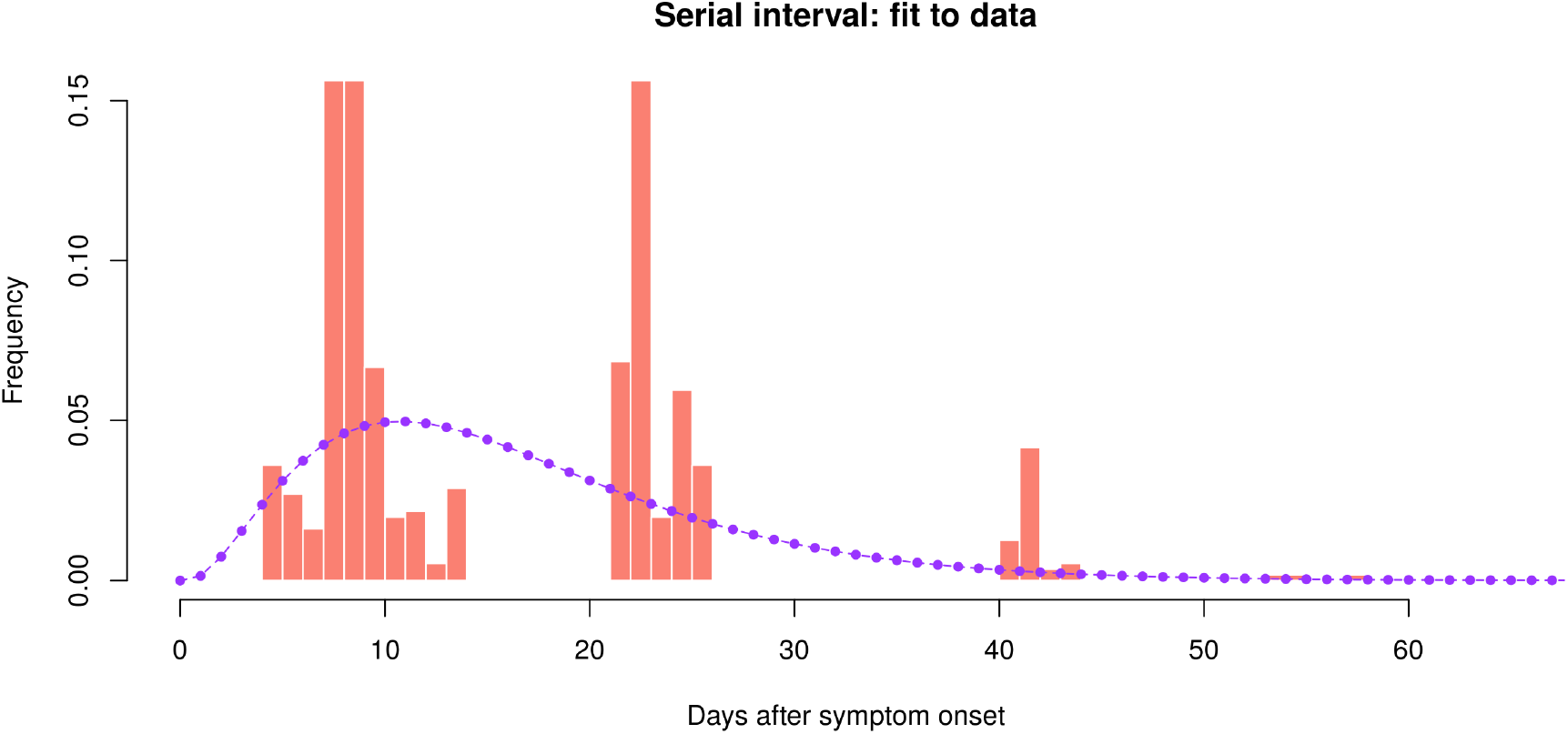
Serial Interval Distribution & Parametric Fit; *µ* = 16.47387; *σ* = 10.18816

**Figure 2.**
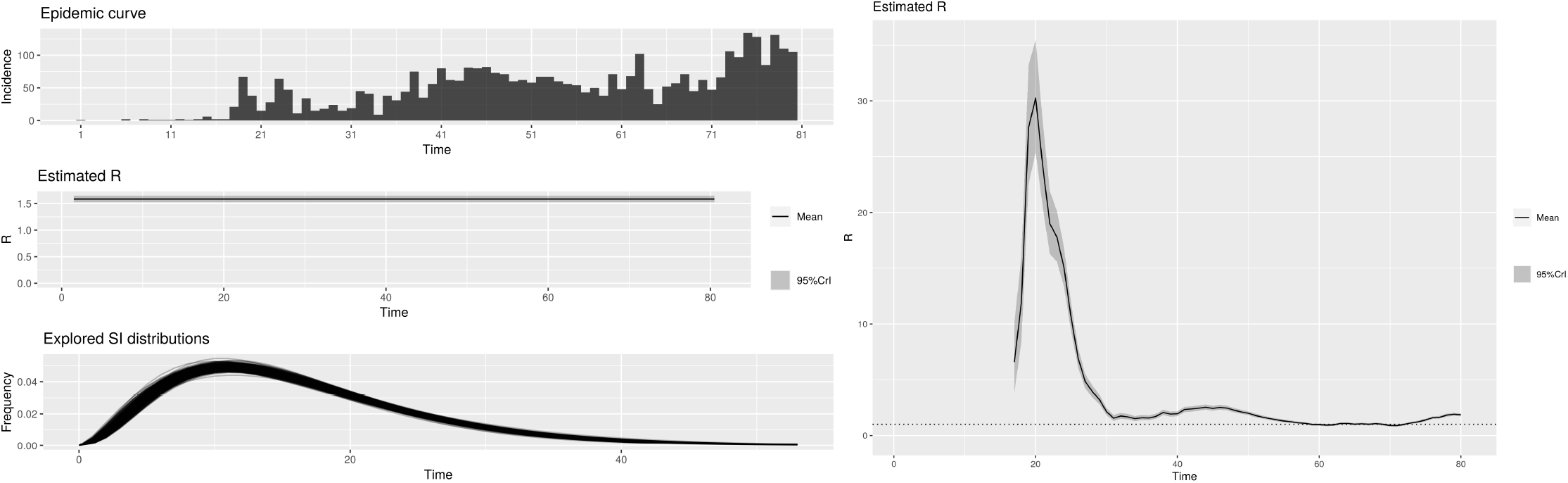
(a) Serial Interval, Incidence, Average Reproduction Number & (b) Instantaneous Reproduction Number, for Andhra Pradesh.

**Figure 3.**
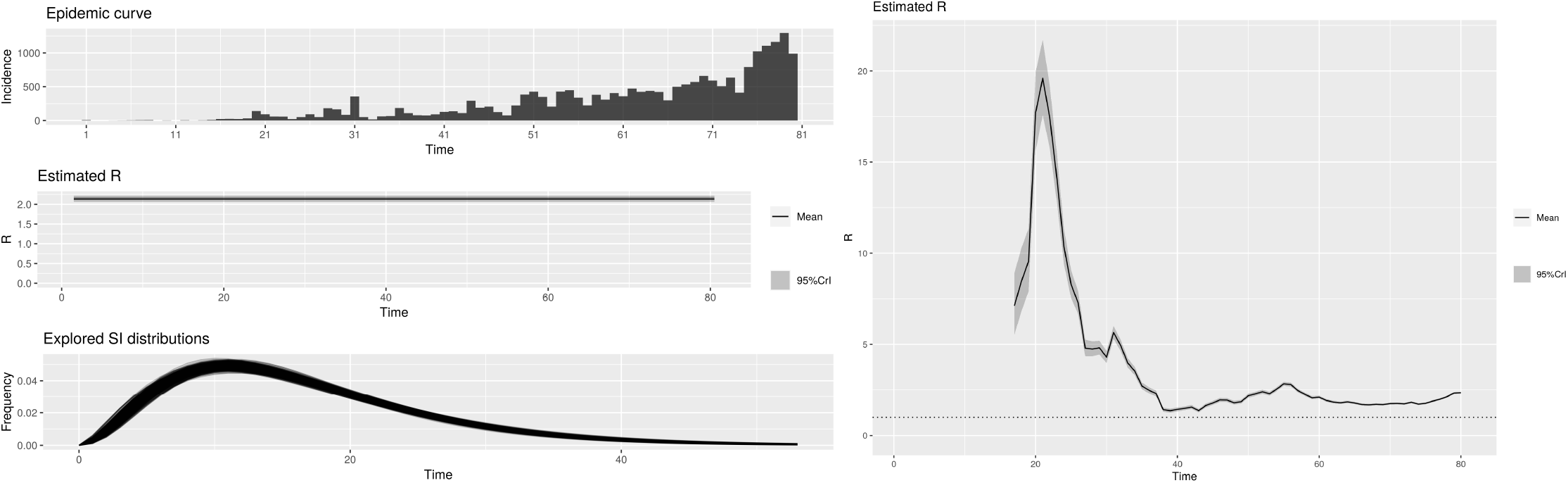
(a) Serial Interval, Incidence, Average Reproduction Number & (b) Instantaneous Reproduction Number, for Delhi.

**Figure 4.**
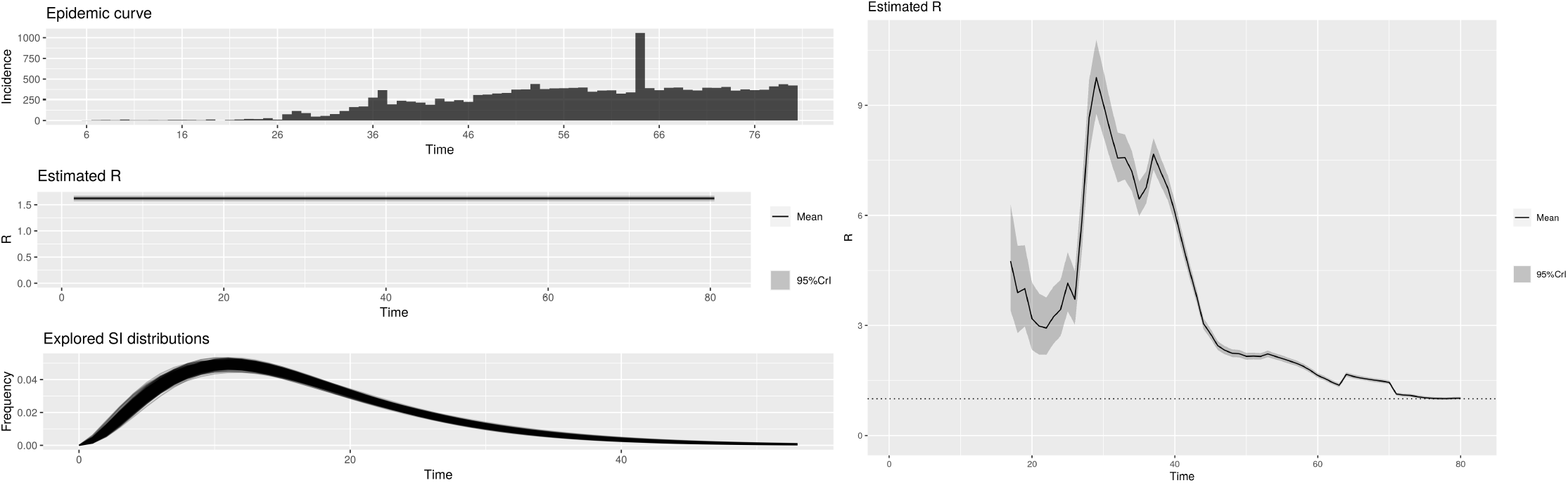
(a) Serial Interval, Incidence, Average Reproduction Number & (b) Instantaneous Reproduction Number, for Gujrat.

**Figure 5.**
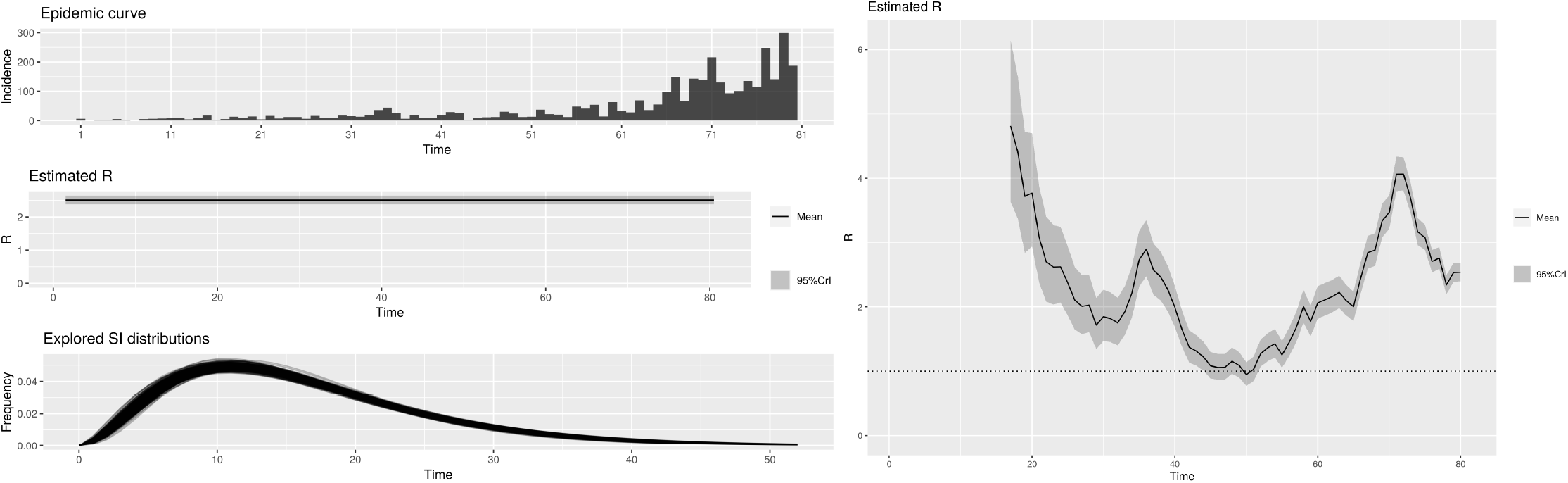
(a) Serial Interval, Incidence, Average Reproduction Number & (b) Instantaneous Reproduction Number, for Karnataka.

**Figure 6.**
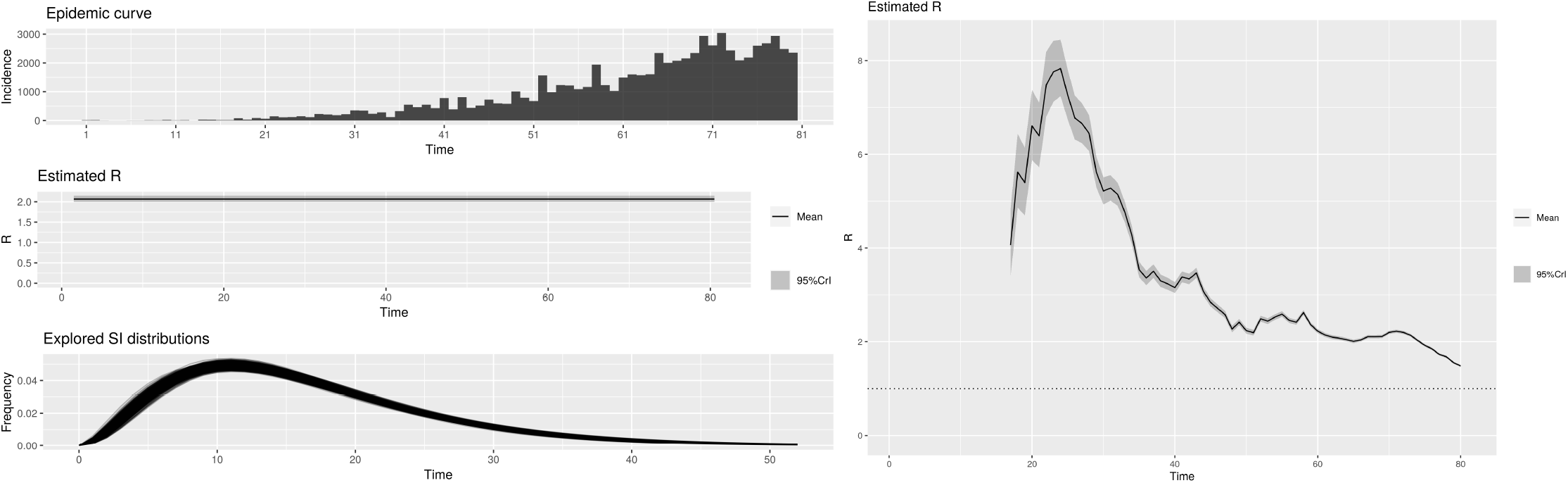
(a) Serial Interval, Incidence, Average Reproduction Number & (b) Instantaneous Reproduction Number, for Maharastra.

**Figure 7.**
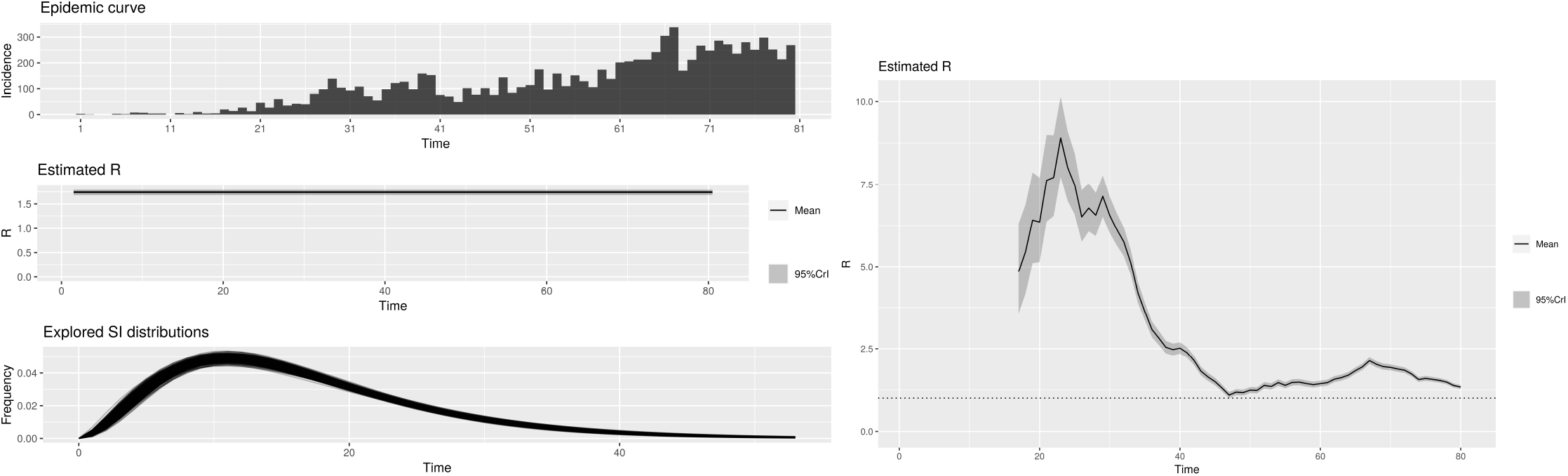
(a) Serial Interval, Incidence, Average Reproduction Number & (b) Instantaneous Reproduction Number, for Rajasthan.

**Figure 8.**
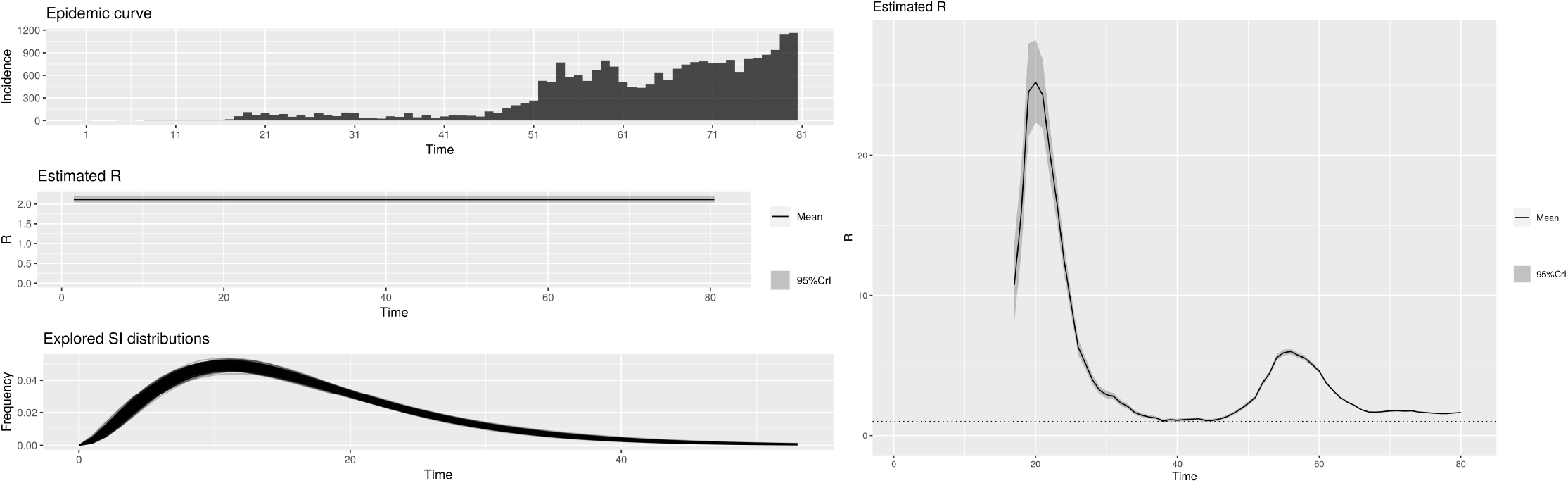
(a) Serial Interval, Incidence, Average Reproduction Number & (b) Instantaneous Reproduction Number, for Tamilnadu.

**Figure 9.**
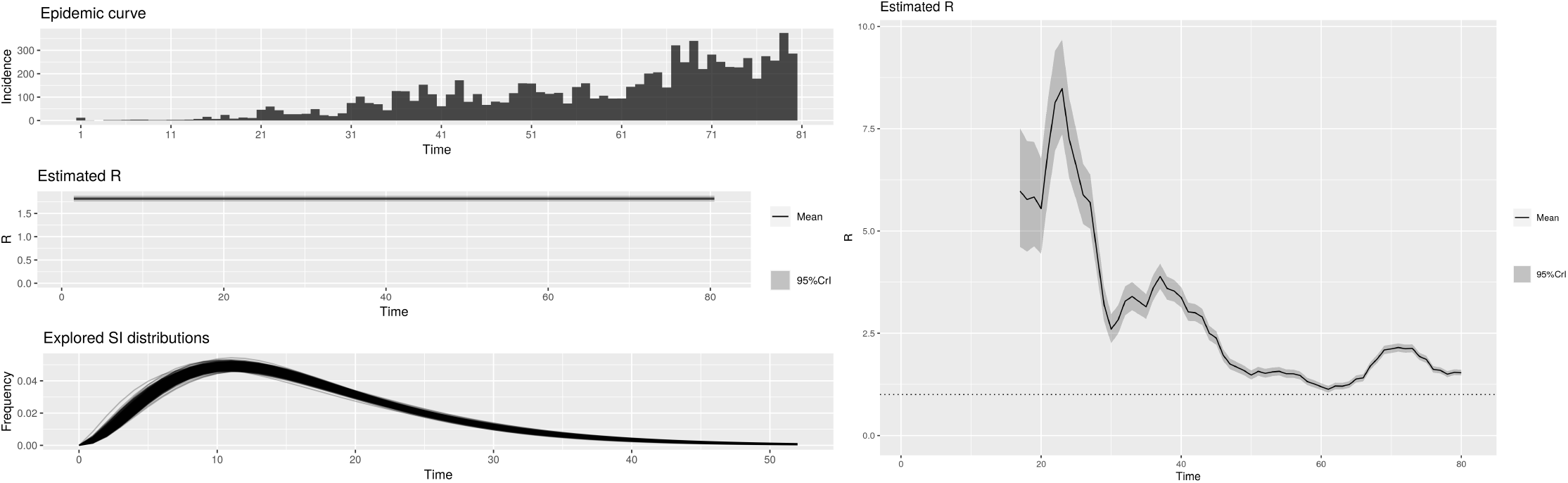
(a) Serial Interval, Incidence, Average Reproduction Number & (b) Instantaneous Reproduction Number, for Uttar Pradesh.

**Figure 10.**
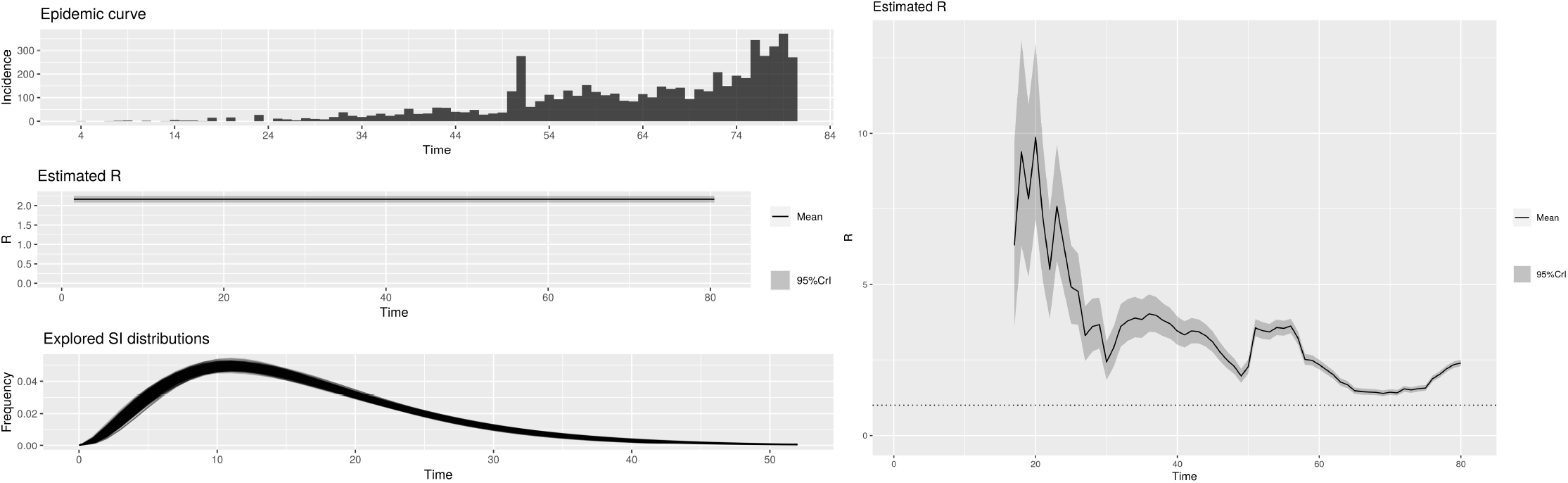
(a) Serial Interval, Incidence, Average Reproduction Number & (b) Instantaneous Reproduction Number, for West Bengal.

**Figure 11.**
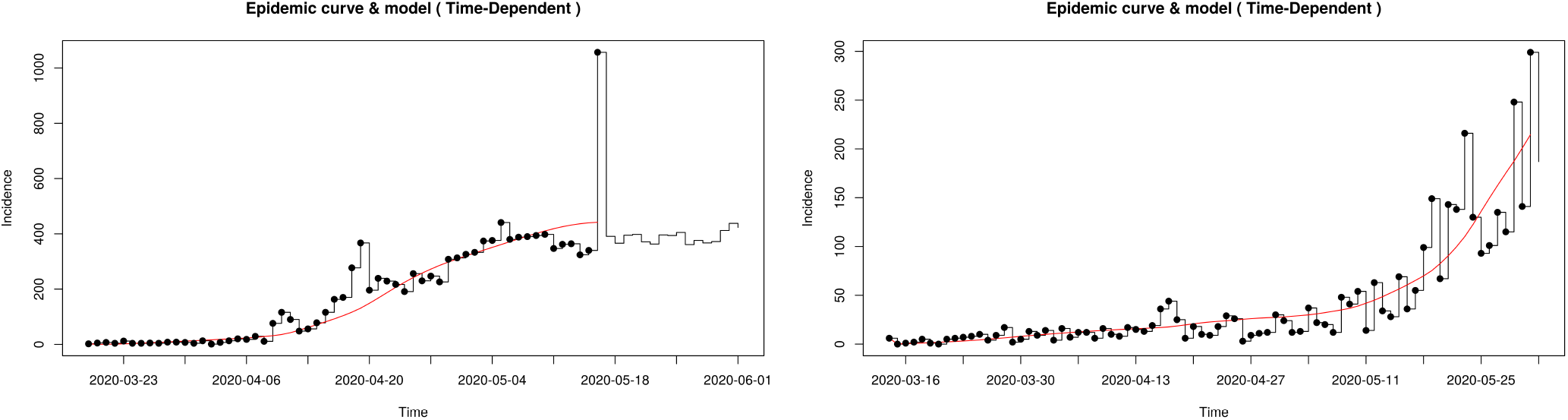
(a) Daily Case Incidences Fit in Gujrat (b) Daily Case Incidences Fit in Karnataka.

## 7. Conclusions

We show that, the lockdown in India was fairly successful barring a couple of places, due to migration or superspreading etc. We note here that a similar study, with a bigger scope has been reported elsewhere [28] But it assumes the parametric serial interval, which is different from ours. We have deduced our own serial interval (cf. sec. 3) by scraping the pan India raw data and by building our own line list & contact list. Hence Our result is presumed to be significantly different from theirs and more representative of the actual scenarios [29]. The effect partial lifting of the lockdown (unlock) is also seen in the results, in terms of increment in R_i_(t).

## Data Availability

If required then we are ready to provide our data on which the article is based to editors / reviewers and independent researchers.

## 8. Acknowledgement

SM wishes to thank K. Bhattacharya, K. Samanta for useful discussions. He also thanks I. Mukhopadhyay for useful help with references. The analysis was was performed in R [30] statistical programming language environment.

